# Deep learning-assisted multiple organ segmentation from whole-body CT images

**DOI:** 10.1101/2023.10.20.23297331

**Authors:** Yazdan Salimi, Isaac Shiri, Zahra Mansouri, Habib Zaidi

## Abstract

**Background:** Automated organ segmentation from computed tomography (CT) images facilitates a number of clinical applications, including clinical diagnosis, monitoring of treatment response, quantification, radiation therapy treatment planning, and radiation dosimetry.

**Purpose:** To develop a novel deep learning framework to generate multi-organ masks from CT images for 23 different body organs.

**Methods:** A dataset consisting of 3106 CT images (649,398 axial 2D CT slices, 13,640 images/segment pairs) and ground-truth manual segmentation from various online available databases were collected. After cropping them to body contour, they were resized, normalized and used to train separate models for 23 organs. Data were split to train (80%) and test (20%) covering all the databases. A Res-UNET model was trained to generate segmentation masks from the input normalized CT images. The model output was converted back to the original dimensions and compared with ground-truth segmentation masks in terms of Dice and Jaccard coefficients. The information about organ positions was implemented during post-processing by providing six anchor organ segmentations as input. Our model was compared with the online available “TotalSegmentator” model through testing our model on their test datasets and their model on our test datasets.

**Results:** The average Dice coefficient before and after post-processing was 84.28% and 83.26% respectively. The average Jaccard index was 76.17 and 70.60 before and after post-processing respectively. Dice coefficients over 90% were achieved for the liver, heart, bones, kidneys, spleen, femur heads, lungs, aorta, eyes, and brain segmentation masks. Post-processing improved the performance in only nine organs. Our model on the TotalSegmentator dataset was better than their models on our dataset in five organs out of 15 common organs and achieved almost similar performance for two organs.

**Conclusions:** The availability of a fast and reliable multi-organ segmentation tool leverages implementation in clinical setting. In this study, we developed deep learning models to segment multiple body organs and compared the performance of our models with different algorithms. Our model was trained on images presenting with large variability emanating from different databases producing acceptable results even in cases with unusual anatomies and pathologies, such as splenomegaly. We recommend using these algorithms for organs providing good performance. One of the main merits of our proposed models is their lightweight nature with an average inference time of 1.67 seconds per case per organ for a total-body CT image, which facilitates their implementation on standard computers.

## Introduction

Segmentation of healthy organs from Computed Tomography (CT) images is critical and beneficial in a number of applications, including the generation of anthropomorphic computational models, delimitation of organs at risk in radiation therapy (RT) treatment planning (1–4), and other kinds of computer-assisted applications, such as pathologic detection (5, 6), prognosis and outcome prediction (7, 8), image quantification (9, 10), and radiation dosimetry calculations (11–13). The manual slice-by-slice segmentation of organs can be labor-intensive and time-consuming, in addition to the high inter- and intra-observer variability reported for segmentation of healthy organs and malignant lesions (14, 15). Since the emergence of machine learning and deep learning (DL) algorithms in medical imaging research, especially medical image segmentation, a number of studies focused on automatic segmentation of structures from CT images and other imaging modalities (16–18). Most published studies attempted to improve segmentation accuracy (commonly quantified by the Dice coefficient), robustness and generalizability on new unseen dataset acquired with different imaging settings on disparate patient characteristics and including a large number of organs (19–21). Newly developed neural network architectures, loss functions, and image processing algorithms contributed to the improvement of the performance of image segmentation models. Yet, the number of datasets and their diversity remains the bottleneck for successful implementation of deep learning-based algorithms (22). Most studies conveyed the performance of the developed models on a test set excluded from the training set, thus reaching very high Dice coefficients as reported in few challenges held on multiple organ segmentations (23). Yet, the majority of these studies didn’t investigate models’ performance on unseen external datasets. Xu et al. (24) focused on the occurrence of outliers during image segmentation and how to solve this problem. Recent studies addressed the limitations and benefits of DL-based organ segmentation in real-life clinical scenarios (14, 25). The comparison of the results achieved by different techniques using private/local databases is not straightforward given that the used datasets are not publicly available. Besides, it’s well established that acquisition, scanner, and demographic parameters can affect the performance of a model on external unseen datasets from other centers (14, 26). Ma et al. (19) described the low performance of segmentation models trained and inferenced on different databases for abdominal organs segmentation task. In this context, a segmentation model trained on a dataset presenting with a large variability and tested on an unseen dataset may be beneficial in estimating the performance in real clinical scenarios.

In this study, we aimed to develop a deep neural network to segment multiple healthy organs (28 organs) from total-body CT images targeting improvement of the accuracy and generalizability compared to previously developed models. We also compared the performance of our models with existing methods and considered the effect of post-processing algorithms to take into account organ-specific anatomical information during the segmentation process.

## Materials and Methods

### Patient population

This study included 3106 CT images (649,398 axial 2D CT slices, 13,640 3D image/segment pairs) collected from multiple online available datasets (27–32). A total of 300 pediatric cases with 18.9 ± 4.13 cm effective diameter and 2806 adult cases with 27.53 ± 5.35 cm effective diameter as defined by the AAPM #204 Report (33) were included. The average age was 6.32 ± 4.34 years for pediatric patients and 66.98 ± 9.84 years for adult patients. It should be noted that the age, gender, and acquisition parameters were available only in a limited number of datasets, the rest were either anonymized or in NIFTI format without additional information. The number of slices and patient size characteristics were summarized in supplementary Table 1. The data were split into training and test set for each organ according to the number of cases from each database to ensure the test and train data use cases from each available database, i.e., the training (80 %) and testing (20%) data for each organ include cases from all databases.

Figure 1 depicts the number of CT images used for training each model for each organ segmentation. The number of training and test datasets are summarized in Table 1. The detailed number of training cases from each database is provided in supplementary Table 2. The masks (segmentations) of the 23 different organs were used to train separate segmentation models. The summed gastrointestinal segment (GIs) was defined by adding distinct segmentations of the duodenum, small intestine, and colon together to define a single organ.

**Figure 1.**
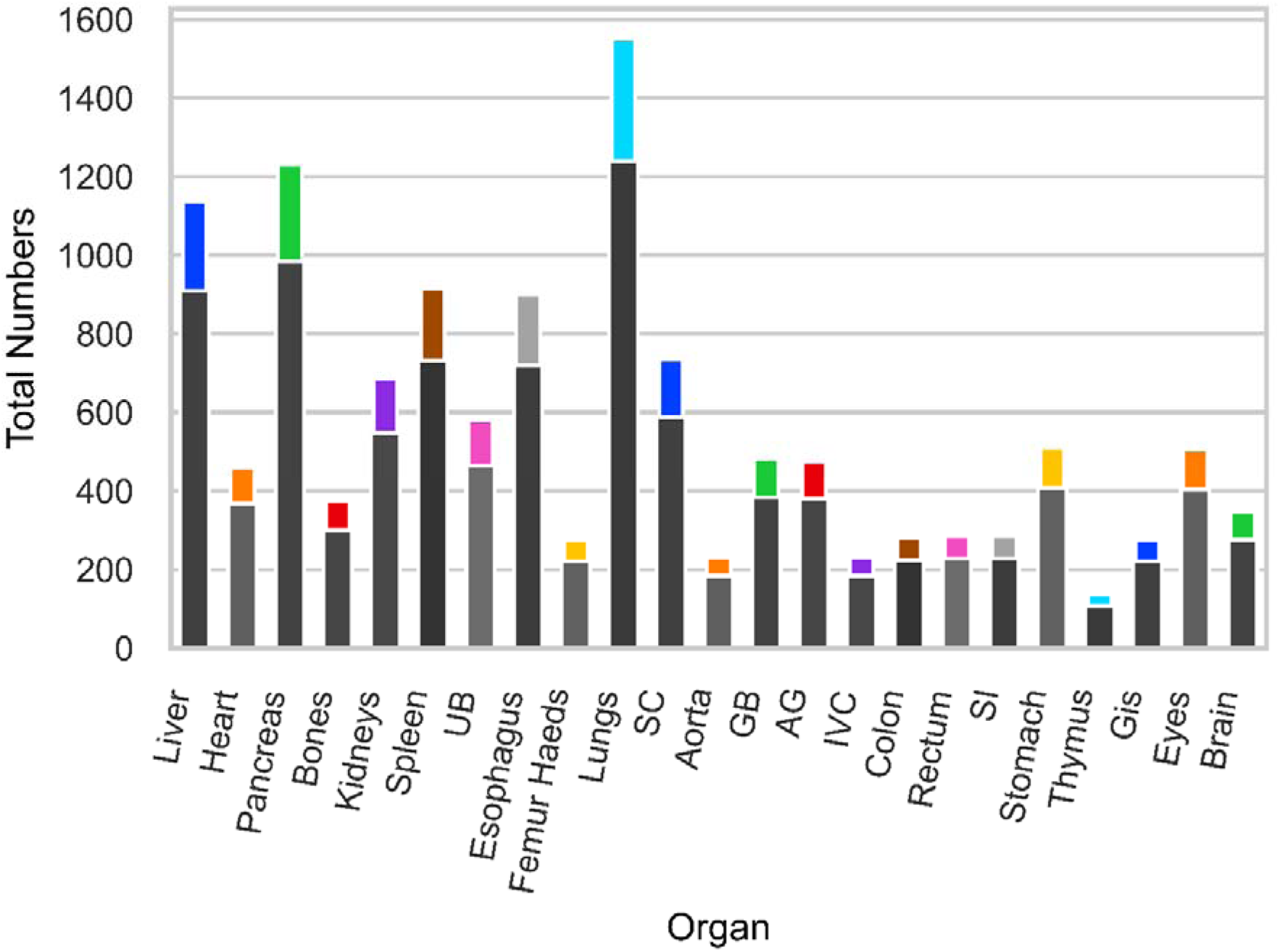
Number of 3D CT images extracted from the clinical studies included in all databases used for training and testing of the models. The upper (color) bar depicts the test, whereas the bottom (gray) bar depicts the training numbers. UB: Urinary Bladder, SC: Spinal Cord, GB: Gall Bladder, AG: Adrenal Gland, IVC: Inferior Vena Cava, SI: Small Intestine, GIs: Gastrointestinal.

**Table 1.** Summary of image segmentation metrics, including Dice, Jaccard coefficients and the effect of post-processing, and the number of cases in the train/validation and test groups in strategy (a). UB: Urinary Bladder, SC: Spinal Cord, GB: Gall Bladder, AG: Adrenal Gland, IVC: Inferior Vena Cava, SI: Small Intestine, GIs: Gastrointestinal.

| Organ | Train# | Test# | Dice_network | Dice_post | Dice_gain_post | Dice_P-value | Jaccard_network | Jaccard_post | Jacc_gain_post | Jacc_P-value |
| --- | --- | --- | --- | --- | --- | --- | --- | --- | --- | --- |
| Liver | 908 | 227 | 96.93 ± 1.67 | 96.98 ± 1.62 | 0.06 ± 0.14 | <0.001 | 94.08 ± 3.01 | 94.19 ± 2.93 | 0.11 ± 0.25 | <0.001 |
| Heart | 368 | 92 | 91.60 ± 6.52 | 91.95 ± 6.13 | 0.35 ± 0.87 | <0.001 | 85.08 ± 9.82 | 85.62 ± 9.38 | 0.54 ± 1.32 | <0.001 |
| Pancreas | 984 | 246 | 77.52 ± 15.99 | 76.24 ± 17.02 | -1.28 ± 5.07 | 0.563 | 65.76 ± 19.15 | 64.25 ± 19.78 | -1.50 ± 5.89 | 0.662 |
| Bones | 300 | 75 | 94.08 ± 4.84 | 93.84 ± 4.75 | -0.24 ± 0.32 | <0.001 | 89.17 ± 7.77 | 88.72 ± 7.60 | -0.45 ± 0.56 | <0.001 |
| Kidneys | 548 | 137 | 94.03 ± 4.80 | 94.13 ± 5.05 | 0.10 ± 0.79 | <0.001 | 89.07 ± 7.57 | 89.28 ± 7.89 | 0.21 ± 1.24 | <0.001 |
| Spleen | 732 | 183 | 94.29 ± 9.88 | 94.68 ± 9.28 | 0.39 ± 3.26 | <0.001 | 90.28 ± 11.81 | 90.85 ± 11.16 | 0.57 ± 4.33 | <0.001 |
| UB | 464 | 116 | 83.41 ± 17.99 | 83.78 ± 17.97 | 0.37 ± 1.34 | <0.001 | 74.75 ± 21.24 | 75.24 ± 21.40 | 0.50 ± 1.84 | <0.001 |
| Esophagus | 720 | 180 | 73.20 ± 10.90 | 72.35 ± 12.55 | -0.84 ± 4.07 | 0.010 | 58.76 ± 12.27 | 58.01 ± 13.73 | -0.75 ± 3.98 | 0.010 |
| Femur Heads | 220 | 55 | 95.65 ± 2.78 | 95.72 ± 2.79 | 0.07 ± 0.14 | <0.001 | 91.78 ± 4.77 | 91.92 ± 4.79 | 0.14 ± 0.24 | <0.001 |
| Lungs | 1240 | 310 | 97.63 ± 1.18 | 96.56 ± 4.70 | -1.07 ± 4.61 | <0.001 | 95.39 ± 2.18 | 93.42 ± 9.06 | -1.97 ± 8.46 | <0.001 |
| SC | 588 | 147 | 89.69 ± 3.65 | 89.85 ± 3.70 | 0.17 ± 0.39 | <0.001 | 81.49 ± 5.63 | 81.76 ± 5.72 | 0.28 ± 0.64 | <0.001 |
| Aorta | 184 | 46 | 92.99 ± 2.85 | 93.55 ± 2.22 | 0.56 ± 1.34 | <0.001 | 87.03 ± 4.77 | 87.95 ± 3.77 | 0.93 ± 2.18 | <0.001 |
| GB | 384 | 96 | 78.74 ± 16.92 | 79.32 ± 16.78 | 0.57 ± 5.35 | 0.095 | 67.60 ± 19.56 | 67.39 ± 21.00 | -0.21 ± 9.09 | 0.298 |
| AG | 380 | 95 | 54.64 ± 17.40 | 51.62 ± 17.52 | -3.02 ± 5.86 | <0.001 | 39.38 ± 15.21 | 30.14 ± 18.41 | -9.24 ± 13.01 | <0.001 |
| IVC | 184 | 46 | 80.57 ± 11.30 | 62.59 ± 11.44 | -17.98 ± 10.78 | <0.001 | 68.77 ± 14.33 | 40.97 ± 13.22 | -27.80 ± 11.26 | <0.001 |
| Colon | 224 | 56 | 70.61 ± 11.97 | 67.48 ± 15.25 | -3.13 ± 7.05 | 0.167 | 55.76 ± 13.20 | 52.16 ± 16.96 | -3.60 ± 8.15 | 0.133 |
| Rectum | 228 | 57 | 77.90 ± 13.31 | 77.70 ± 13.33 | -0.20 ± 2.44 | 0.064 | 65.41 ± 15.13 | 65.14 ± 15.23 | -0.26 ± 3.31 | 0.048 |
| SI | 228 | 57 | 70.79 ± 14.23 | 71.45 ± 14.52 | 0.66 ± 1.00 | <0.001 | 56.35 ± 14.63 | 56.67 ± 16.10 | 0.32 ± 4.34 | <0.001 |
| Stomach | 408 | 102 | 86.22 ± 10.76 | 75.13 ± 12.17 | -11.09 ± 11.46 | <0.001 | 77.12 ± 14.36 | 57.20 ± 18.80 | -19.92 ± 18.09 | <0.001 |
| Thymus | 108 | 27 | 64.11 ± 23.55 | 65.36 ± 23.57 | 1.25 ± 2.63 | <0.001 | 50.89 ± 22.67 | 52.26 ± 22.75 | 1.37 ± 3.01 | 0.002 |
| Gis | 220 | 55 | 88.88 ± 8.87 | 89.26 ± 8.22 | 0.38 ± 1.44 | <0.001 | 80.91 ± 11.79 | 81.40 ± 11.14 | 0.50 ± 1.66 | <0.001 |
| Eyes | 404 | 101 | 91.13 ± 3.13 | 91.13 ± 3.15 | 0.00 ± 0.09 | 0.860 | 83.84 ± 5.09 | 25.75 ± 38.89 | -58.09 ± 39.10 | <0.001 |
| Brain | 276 | 69 | 97.39 ± 1.66 | 96.72 ± 5.26 | -0.67 ± 4.48 | 0.909 | 94.95 ± 3.03 | 33.10 ± 44.58 | -61.85 ± 44.10 | <0.001 |

### Preprocessing and Network architecture

The external body contour was extracted from axial CT images through image processing algorithms developed and used in previous studies (34, 35). The CT images were cropped to a bounding box (BB) including the body contour in the lateral and AP directions to remove the background area. The images were cropped to 30 slices in the superior direction and 30 slices in the inferior direction according to the BB covering organ segmentation in the Z-axis (cranio-caudal direction). The model was trained in 2D fashion, meaning that the input to the network consisted of 2D axial images with the output being the corresponding segmentation masks. A Res-UNET neural network architecture used in a previous PET segmentation study (36) was employed in this work (Figure 2). The 2D images and masks were resized to 304 (right to left) × 224 (anterior posterior) pixel dimensions. The image intensities were clipped between −70 HU and +170 HU and normalized between zero and one and then discretized to 240 intensity values.

**Figure 2.**
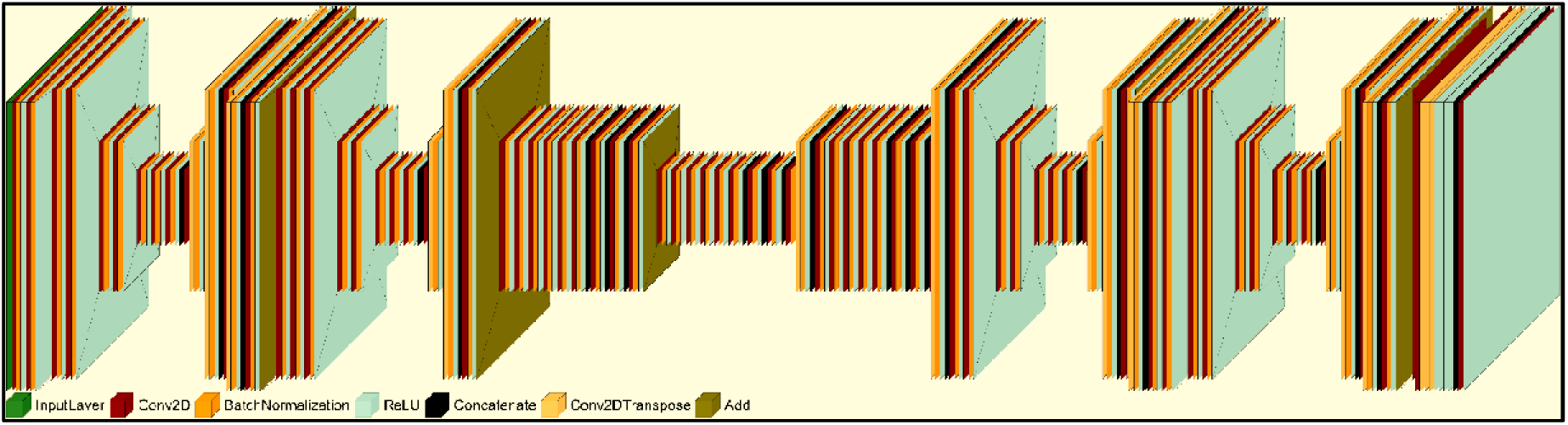
The Res-UNET neural network architecture adopted for multi-organ segmentation from CT images. Conv2d: 2D convolution layer.

The cropped information was stored in the image header and used later to reverse the cropped model output segmentation to the original image dimensions. Figure 3 summarizes the steps performed to train the network.

**Figure 3.**
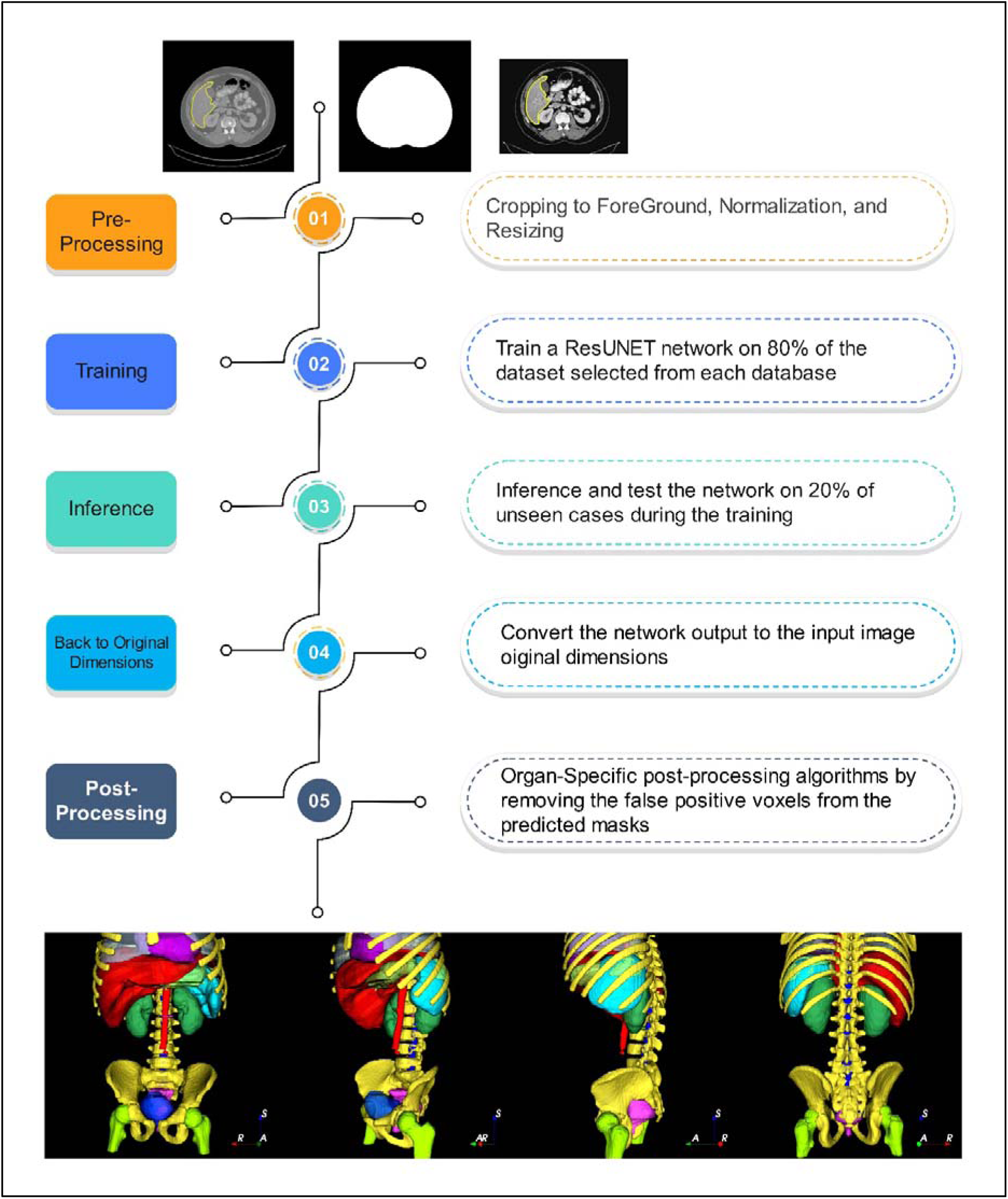
The flowchart of training and inference steps followed in implementing the segmentation algorithm. From left to right: The upper axial slices show CT images, with the liver mask defined with the yellow contour, segmented body contour, and pre-processed (cropped, normalized, and resized) images. The lower segmentation shows the 3D visualization of the network output after post-processing.

### Post-processing and prior knowledge implementation

Organ-specific post-processing algorithms were used to take into account the anatomical locations of the organs. For all test datasets, segmentation masks of six anchor organs including the liver, spleen, lung, femoral head, bladder, and kidneys were generated by our trained models. These generated segmentation masks were used to perform organ-specific post-processing. For each organ, a specific algorithm was used to remove the segmented voxels outside of the BB delimitating each organ. For instance, for spleen post-processing, the post-processing function input was the liver, lung, and femoral head segmentation. The BB of the femur and lung were determined considering the known prior anatomical information that the liver is in the abdomen and always higher than the femoral head BB and lower than the lungs apex. We used the same strategy for other organs, such as the gall bladder (GB), and adrenal glands (AG) to remove unwanted (false positive) voxels from the network output by exploiting the fact that the gall bladder is at the inferior part of the liver and the adrenal gland are upper than kidneys. In the end, the network performance evaluation was compared with/without organ-specific post-processing.

### Evaluation metrics

For each organ segmentation task, 20% of each database was randomly defined and used as the test dataset. The predicted segmentation masks were compared to the ground truth masks by measuring Dice and Jaccard coefficients.

### Real-life evaluation on external datasets

To evaluate the performance of our model in real clinical scenarios on an external unseen dataset and compare our method to previously reported deep learning models, we tested our trained models on the online available TotalSegmentator dataset published recently by Wasserthal et al. (37) and then tested their trained model on the databases we used for testing. The dataset used in the above reference was local for the involved centers and could be considered completely unseen data for our models, In addition, they separated the test and train dataset to make the comparison more reproducible. They have used state-of-the-art nnU-Net (38) network/training methodology and managed to won 9 out of 10 MICCAI 2020 (39) and AMOS (23) challenges. We compared the performance of our model and models reported in the reference above for organs included in both studies (15 organs listed in Table ***2***). Figure 4 shows the dataflow in this study. Overall, we performed three evaluations: a) our model tested on our test dataset (23 organs), b) our model tested on the TotalSegmentator test dataset (15 organs), and c) the TotalSegmentator model tested on our dataset (15 organs). TotalSegmentator trained model was collected from GitHub on April 28, 2023.

**Figure 4.**
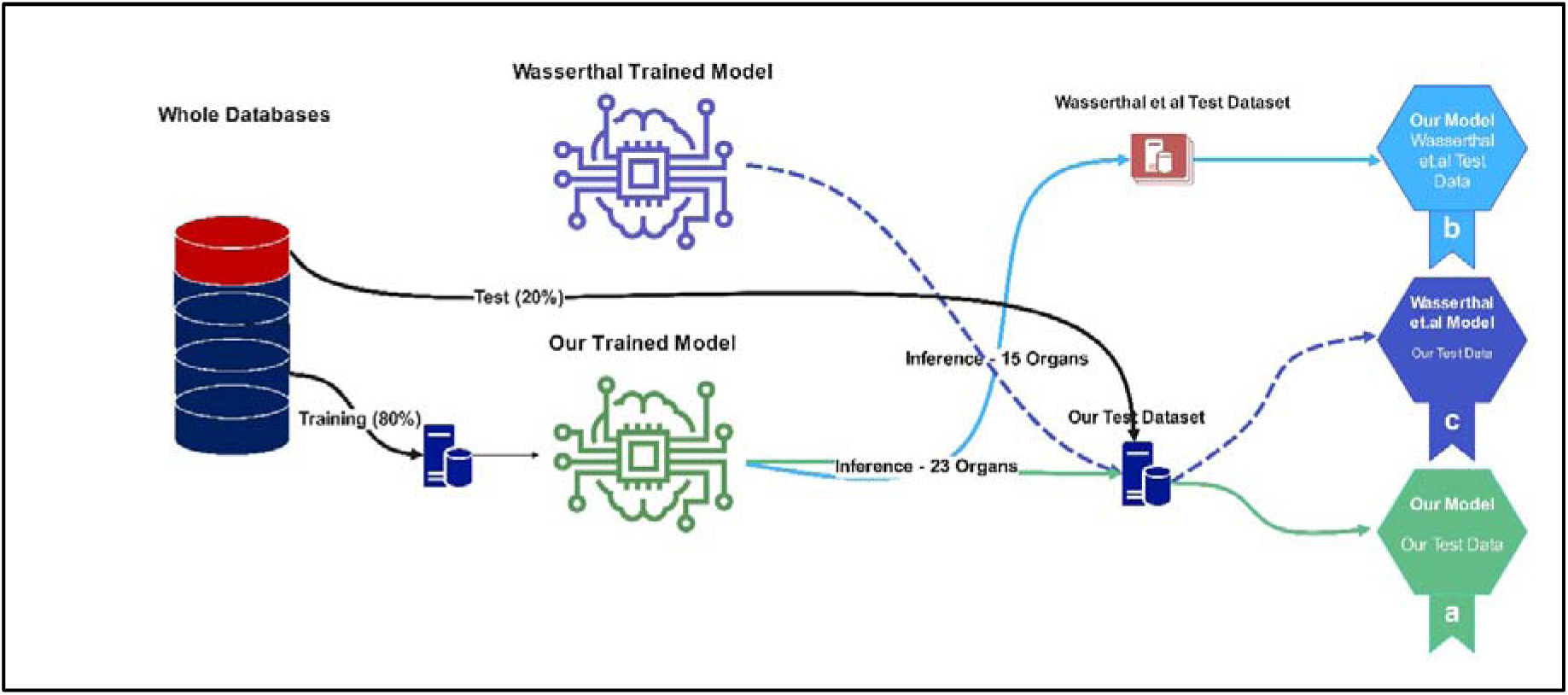
The Dataflow adopted for external evaluation. Green, light blue, and dark blue lines show the dataflow for strategies (a), (b), and (c), respectively.

**Table 2.** Results of the external evaluation using Wasserthal et al. (37) algorithm in strategies (b) and (c). * The brain images were distorted and blurred in Wasserthal et al. dataset. UB: Urinary Bladder, SC: Spinal Cord, GB: Gall Bladder, AG: Adrenal Gland, IVC: Inferior Vena Cava, SI: Small Intestine, GIs: Gastrointestinal.

| Organ | Wasserthal et al. model on our test data (c) |  |  | Our model on Wasserthal et al. test data (b) |  |  |  |  |
| --- | --- | --- | --- | --- | --- | --- | --- | --- |
|  | Test # | Dice_summary_test | Jaccard_summary_test | Test # | Dice_network | Dice_post | Jaccard_network | Jaccard_post |
| Liver | 227 | 87.18 ± 27.58 | 83.87 ± 27.11 | 46 | 94.96 ± 3.57 | 95.02 ± 3.67 | 90.60 ± 6.11 | 87.40 ± 16.82 |
| Pancreas | 246 | 57.61 ± 35.31 | 48.04 ± 30.94 | 37 | 70.36 ± 19.36 | 68.60 ± 19.31 | 57.05 ± 19.35 | 29.73 ± 24.94 |
| Kidneys | 137 | 91.44 ± 6.34 | 84.76 ± 9.36 | 39 | 88.47 ± 7.60 | 90.41 ± 7.56 | 80.03 ± 10.75 | 79.44 ± 21.59 |
| Spleen | 183 | 90.38 ± 19.37 | 85.84 ± 19.51 | 45 | 93.13 ± 6.31 | 93.68 ± 4.97 | 87.73 ± 10.00 | 85.57 ± 15.39 |
| UB | 116 | 75.03 ± 26.66 | 65.63 ± 26.79 | 36 | 77.91 ± 18.06 | 80.17 ± 17.18 | 66.67 ± 20.29 | 65.25 ± 26.47 |
| Esophagus | 180 | 72.25 ± 13.46 | 58.05 ± 14.36 | 44 | 51.75 ± 14.69 | 53.42 ± 17.00 | 36.18 ± 13.22 | 24.86 ± 26.43 |
| Femur Heads | 55 | 81.15 ± 17.30 | 70.94 ± 18.91 | 30 | 79.24 ± 12.72 | 86.52 ± 10.44 | 67.30 ± 16.56 | 71.41 ± 25.32 |
| Lungs | 310 | 97.81 ± 1.10 | 95.74 ± 2.04 | 47 | 96.64 ± 2.33 | 96.33 ± 3.53 | 93.60 ± 4.23 | 85.06 ± 26.90 |
| Aorta | 46 | 90.98 ± 2.57 | 83.56 ± 4.26 | 46 | 82.01 ± 7.85 | 82.53 ± 10.37 | 70.20 ± 10.64 | 69.90 ± 16.12 |
| GB | 96 | 77.63 ± 17.72 | 66.07 ± 18.74 | 31 | 69.72 ± 25.72 | 72.59 ± 26.15 | 58.24 ± 25.09 | 57.86 ± 30.36 |
| AG | 95 | 55.46 ± 20.49 | 41.07 ± 19.40 | 36 | 44.12 ± 21.95 | 43.98 ± 22.10 | 30.72 ± 17.78 | 15.95 ± 16.57 |
| Colon | 56 | 65.55 ± 14.22 | 50.28 ± 14.83 | 42 | 46.83 ± 19.88 | 45.28 ± 18.82 | 32.59 ± 16.12 | 27.67 ± 17.78 |
| SI | 57 | 58.85 ± 16.82 | 43.63 ± 16.71 | 39 | 39.62 ± 17.72 | 41.10 ± 18.38 | 26.24 ± 14.34 | 25.79 ± 17.11 |
| Stomach | 102 | 88.59 ± 11.43 | 80.80 ± 12.86 | 46 | 59.42 ± 26.78 | 58.14 ± 25.73 | 46.95 ± 25.49 | 22.45 ± 17.98 |
| Brain* | 69 | 90.13 ± 17.80 | 85.29 ± 20.51 | 9 | 66.52 ± 37.45 | 65.51 ± 36.75 | 58.62 ± 35.63 | 35.59 ± 42.47 |

### Statistical analysis

We used the Wilcoxon rank t-test to evaluate the effect of post-processing on each organ.

## Results

### Evaluation on the test dataset

Table 1 summarizes the Dice and Jaccard image segmentation metrics for our model on test sets separated from our data (strategy (a) mentioned in the *Methods* section). The highest Dice coefficients were achieved for the lung, spleen, liver, and brain organs, while the lowest values were obtained for the thymus, adrenal gland organs. Organ-specific post-processing increased the Dice coefficient in seven organs by more than 0.15 absolute value and this increase was statistically significant. However, it did not increase or even significantly decrease the Dice coefficient for the remaining organs. These seven organs were the heart, spleen, UB, SC, aorta, GB, and thymus. Figure 5 depicts examples of 3D visualization of segmentations of CT images corresponding to different subjects shown from eight different perspectives. Supplementary figure 1 extends the examples shown in Figure 5 for pediatric cases and cases with unusual anatomical variations such as splenomegaly. Figure 6 depicts the Dice coefficients in different organs with/without post-processing. The detailed results of segmentation accuracy for each database included in the assessment are presented in supplementary Table 2.

**Figure 5.**
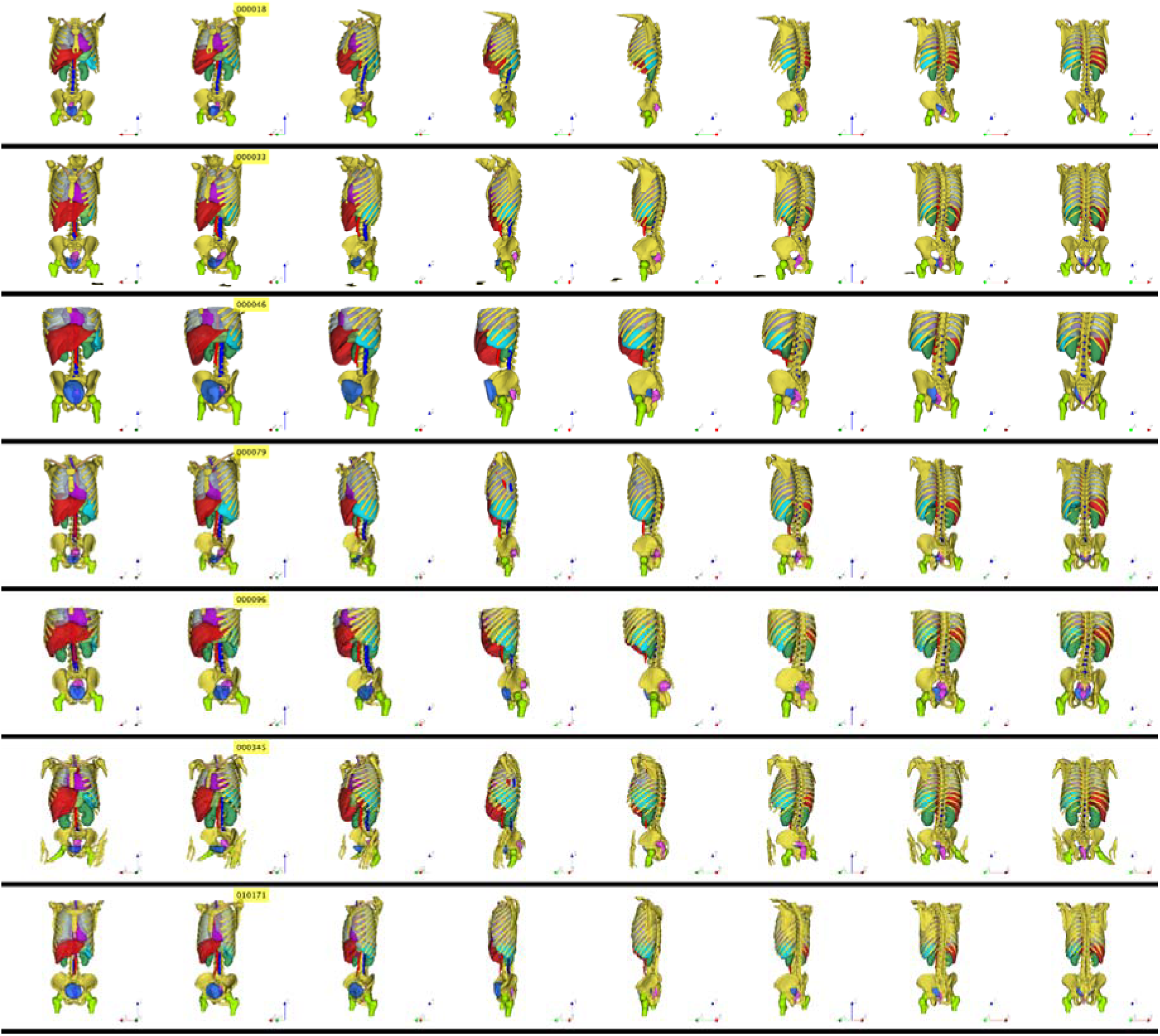
3D visualization of segmentation masks of different organs showing: the kidneys (dark green), femoral heads (lime), bones (yellow), liver (dark red), aorta (light red), spleen (cyan), heart (purple), stomach (light green), spinal cord (dark blue), urinary bladder (light blue), and the rectum (pink).

**Figure 6.**
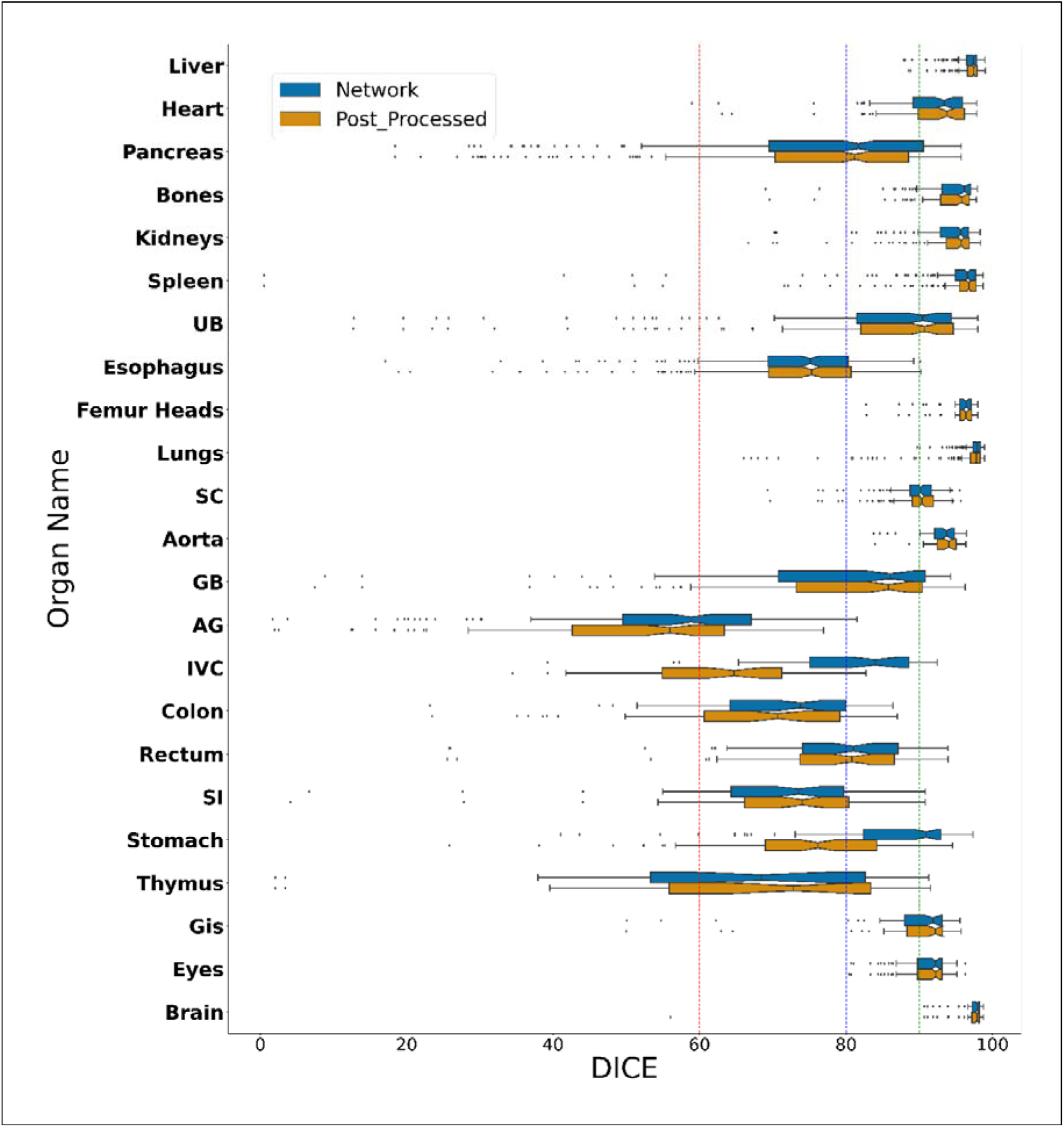
Box plots of Dice coefficients achieved for different organ segmentations before and after post-processing. The red, blue, and green reference lines depict 60%, 80%, and 90% Dice coefficients, respectively. UB: Urinary Bladder, SC: Spinal Cord, GB: Gall Bladder, AG: Adrenal Gland, IVC: Inferior Vena Cava, SI: Small Intestine, GIs: Gastrointestinal.

### External comparison

Table 2 summarizes the Dice and Jaccard evaluation metrics for strategies (b) and (c). A relative difference larger than 2% for the Dice coefficient was considered as significant when comparing strategies (b) and (c). Our model outperformed TotalSegmentator for five organs, including the liver, pancreas, spleen, UB, and femur heads, while the outcome was the same for the lungs and kidneys. For the remaining organs listed in Table ***2*** (7 organs), the TotalSegmentator models outperformed our models. The brain mask was not valid due to the blurring generated on the face area for privacy preserving concerns on TotalSegmentator dataset. Organ-specific post-processing improved the segmentation accuracy, reflected by higher Dice coefficients for strategy (b) in seven other organs, including the liver, kidneys, spleen, UB, esophagus, femur heads, and GB. Table 3 compares our model’s performance in strategy (a) to recent studies reported in the literature for common organs.

**Table 3.** Model performance reported in terms of Dice coefficient in strategy (a) compared to recent studies in the field. UB: Urinary Bladder, SC: Spinal Cord, GB: Gall Bladder, AG: Adrenal Gland, IVC: Inferior Vena Cava.

| Study | Liver | Heart | Pancreas | Kidneys | Spleen | UB | Esophagus | Femur Heads | Lungs | SC | Aorta | GB | AG | IVC | Colon | Rectum | Stomach | Eyes | Brain |
| --- | --- | --- | --- | --- | --- | --- | --- | --- | --- | --- | --- | --- | --- | --- | --- | --- | --- | --- | --- |
| this study | 96.98 | 91.95 | 77.52 | 94.13 | 94.68 | 83.78 | 73.20 | 95.72 | 97.63 | 89.85 | 93.55 | 79.32 | 54.62 | 80.57 | 70.60 | 77.90 | 86.22 | 91.13 | 97.39 |
| ang et al. (40) | 96.60 |  | 76.00 | 93.80 | 96.30 |  | 78.80 |  |  |  | 92.30 | 82.60 | 73.60 | 85.30 |  |  | 85.70 |  |  |
| uan et al. (41) |  |  |  |  |  | 93.00 |  | 96.00 |  |  |  |  |  |  |  | 85.00 |  |  |  |
| jiao et al. (42) | 96.00 |  |  | 84.00 |  | 92.00 |  | 95.00 |  |  | 90.00 | 83.00 |  | 78.00 |  | 83.00 | 89.00 |  |  |
| ong et al. (43) | 95.35 |  | 74.81 |  | 94.07 | 83.81 | 78.26 |  |  |  |  |  |  |  |  |  | 89.01 |  |  |
| ang et al. (44) | 96.76 |  | 81.22 | 92.57 (L)<br>88.06 (R) | 84.21 |  |  |  |  |  | 90.76 |  |  |  |  |  | 80.93 |  |  |
| respi et al. (45) |  | 93.20 |  |  |  |  | 75.90 |  | 96.98 (R)<br>97.36 (L) | 89.42 |  |  |  |  |  |  |  |  |  |
| hi et al. (1) | 98.00 | 96.90 | 90.70 | 97.90 | 96.90 | 95.50 (Male)<br>90.20 (Female) | 97.50 | 98.10 | 98.80 | 91.10 | 93.40 | 94.40 |  |  | 87.40 | 93.70 | 97.80 | 97.70 (R)<br>97.20 (L) | 99.30 |
| fa et al. (19) |  |  |  |  |  |  |  |  |  |  |  |  |  |  |  |  |  |  |  |
| hao et al. (21) | 95.47 |  | 74.52 | 95.49 | 96.15 |  | 76.92 |  |  |  |  | 84.79 |  |  |  |  | 90.21 |  |  |
| iciarz et al. (46) |  |  |  |  |  |  | 84.00 |  |  | 86.00 |  |  |  |  |  |  |  | 90.00 (R)<br>92.00 (L) | 97.00 |
| hen et al. (47) | 95.56 | 93.88 |  | 94.02 |  |  | 81.60 |  | 99.33 |  |  |  |  |  |  |  | 89.15 |  |  |

## Discussion

Automated organ segmentation is a critical step in a wide range of clinical applications, including personalized radiation dosimetry and quantification, and radiation treatment planning. The availability of a fast and reliable organ segmentation tool can facilitate the automation of these procedures and their adoption/deployment in clinical setting. In this work, we developed novel deep learning models to segment multiple organs from total-body CT images and compared the performance of our models with previous algorithms reported in the literature. Our model was trained on images presenting with high variability using large datasets, including adults, pediatrics and patients presenting with a wide range of pathologies and anatomic pathologies. The proposed model demonstrated an acceptable outcome even on cases with uncommon anatomies and pathologies, such as splenomegaly as shown in **Error! Reference source not found.Error! Reference source not found.**. Besides, we used prior anatomical knowledge for some organs in the body for organ-specific post-processing to improve the outcome. This methodology enabled to successfully improve the results in nine organs by achieving higher Dice coefficients in strategy (a) and could help improving the Dice coefficient in more than five organs in strategy (b). We used the model output anchor organ segmentation as reference for prior knowledge implementation i.e., no manual segmentation or ground truth segmentation was needed in post-processing. The error in anchor organ masks was used as input for the post-processing function that can propagate to the post-processed mask and accumulated errors can be problematic. One possible explanation is that our decision algorithm to exclude false positive segmented voxels was not successful in a number of organs. We believe that using ground truth anchor organ segmentations for post-processing can improve the post-processing capabilities. According to the results achieved through post-processing, we suggest using these algorithms for organs achieving good performance (Table ***2*** and Table 3).

To evaluate the performance of the proposed model on real-world unseen external datasets, we tested our model on the online available dataset provided by Wasserthal et al. (37). Our model outperformed their model for five organs in strategies (b) and (c). For most small organs and gastrointestinal organs, our model’s performance was inferior to their model, which can be explained by the different 2D and 3D training strategies used in our and their models, respectively. They have used the nn-Unet (38) model which demands a high computational burden for both training and inference. Their images were resampled to 1.5 mm isotropic voxel dimension in a specific orientation. Conversely, we resampled the images again during cropping and resizing in strategy (b), while our test set in strategy (c), the images were in the original dimensions and the used preprocessing steps were similar to the training step in their model. As shown in

Table 2, the number of valid cases in the testing group was limited and the effect of statistical difference can be significant, while the number of our test dataset was larger. It should be mentioned that their brain images were blurred in the face region to preserve the privacy of subjects. This has affected the performance of our model in strategy (b). In addition, their initial model was trained and improved based on models trained on the same dataset used for training our models and then manually edited the segmentations. As such, our test dataset was not really unseen for their model in strategy (c), while in strategy (b) their local data were completely unseen for our model. The effect of post-processing improved the performance of image segmentation of five organs and there is still scope for improvement that can be explored in future studies by changing the number of anchor organs or providing manual edited segmentation as an aid to generate robust post-processing without initial anchor segmentation errors.

The comparison of our models with recent studies revealed that our model’s performance was better or at least comparable to algorithms reported in the literature for most organs, except for small and gastrointestinal organs (Table 3). One of the main merits of our proposed model is its lightweight requiring a small number of parameters (533 K). In addition, for each organ, we have a separate light model, and the user can select a lower number of organs to be segmented to save time. We calculated the inference time on an NVIDIA RTX 4090 GPU where the average inference time for a total body CT was 1.67 seconds per case per organ. Besides, we tested inferring our model on an Intel Corei9 13900KF CPU and the inference time on the CPU was 14.5 seconds per study and per organ, which is a bearable and acceptable inference time for centers lacking access to dedicated GPUs.

We trained different deep learning models to segment 23 organs from total-body CT images which can be beneficial in various clinical tasks. We evaluated our models on an external dataset. The number of cases was limited to a few organs, and the segmentation criteria were different for each manual segmentation available from the online databases, inherently causing inter-observer variability, e.g., some databases provided whole segmented kidneys while others excluded pelvicalyceal systems. These differences may mislead our models and affect their performance.

## Conclusion

We have developed a fully automated deep learning algorithm capable of generating accurate masks for multiple organs from CT images in an affordable computing time. After training these models on a diverse dataset comprising images from various databases, we compared the performance of our model with other algorithms on external datasets in real clinical scenarios. The proposed model exhibited remarkable capabilities even in cases involving uncommon anatomies and pathologies, such as splenomegaly.

Based on our analysis and results, we recommend using this algorithm especially for organs achieving excellent performance. One key advantage of our proposed models is their lightweight nature, enabling to run them efficiently on standard devices without access to dedicated GPUs in a bearable time. With an average GPU inference time of only 1.67 seconds per organ for a total-body CT image, they provide fast results and can be exploited in most routine tasks, even for verification in RT positioning. Overall, a reliable organ segmentation tool enables wider adoption by medical professionals in clinical setting.

## Data Availability

Dataset is available on request

## Acknowledgments

This work was supported by the Euratom research and training programme 2019-2020 Sinfonia project under grant agreement No 945196.

