## Supplementary material for "Deep learning-assisted multiple organ segmentation from whole-body CT images": Supplemntary File

**Short running title:** Deep learning-assisted multiple organ segmentation

**Supplementary Table 1.** Summary of the number of CT slices and patient size metrics. Datasets of #18 and #19 was mostly cropped to a region partially covering the body, so the DW and diameters were not included.

| Database code | Number of CT slices | DW (cm) | AP diameter (cm) | Lateral diameter (cm) | Effective diameter (cm) |
| --- | --- | --- | --- | --- | --- |
| Database #0 | 130.35±67.5 | 25.41±2.37 | 20.58±2.58 | 32.09±2.54 | 25.87±2.48 |
| database #1 | 155.1±40.96 | 29.5±3.67 | 23.82±4.18 | 40.07±5.44 | 30.02±4 |
| database #2 | 141.38±62.55 | 21.5±2.51 | 19.45±1.61 | 26.31±4.41 | 21.34±2.95 |
| database #3 | 456.18±268.91 | 30.1±4.72 | 25.84±4.95 | 36.79±4.52 | 31.05±4.93 |
| database #4 | 126.48±76.39 | 29.51±3.39 | 25.82±3.57 | 34.71±3.77 | 30.38±3.6 |
| database #5 | 92.22±11.20 | 19.69±1.77 | 18.79±1.44 | 23.18±2.78 | 19.27±1.82 |
| database #6 | 94.56±8.49 | 16.55±0 | 16.4±0 | 18.17±0 | 16.06±1.22 |
| database #7 | 135.94±13.45 | 20.72±1.54 | 18.6±1.22 | 26.24±2.01 | 20.66±1.59 |
| database #8 | 147.26±20.77 | 21.54±2.06 | 19.38±1.65 | 26.61±3.03 | 21.49±2.2 |
| database #9 | 159.51±31.99 | 27.44±3.58 | 23.27±3.17 | 38.9±3.75 | 28.98±3.56 |
| database #10 | 121.39±23.42 | 26.58±3.01 | 22.67±2.56 | 36.74±3.31 | 28.57±2.92 |
| database #11 | 180.23±18.92 | 21.26±2.04 | 19.32±2.14 | 26.73±2.4 | 21.29±2.17 |
| database #12 | 236.78±49.41 | 29.39±3.45 | 25.81±3.81 | 34.27±3.66 | 30.25±3.74 |
| database #13 | 275.72±215.04 | 18.78±3.96 | 15.34±3.34 | 23.39±5.12 | 18.93±4.13 |
| database #14 | 128.2±39.86 | 21.58±1.87 | 19.57±1.32 | 26.7±3.69 | 21.78±2.06 |
| database #15 | 447.62±275.25 | 26.97±5.29 | 26.09±4.82 | 36.82±4.54 | 31.29±4.8 |
| database #16 | 95.09±48.47 | 27.77±3.55 | 25.82±3.52 | 34.8±3.79 | 30.41±3.58 |
| database #17 | 89.02±36.75 | 28.64±4.54 | 27.25±4.25 | 36.92±4.43 | 32±4.17 |
| database #18 | 212.64±194.44 | NA | NA | NA | NA |
| database #19 | 537.98±359.36 | NA | NA | NA | NA |

**Supplementary Table2.** Summary of the number of cases from the different databases enrolled in the study (train and test), impact of post-processing on the achieved outcomes in terms of Dice and Jacquard coefficients.

| **Organ** | **Database** | **Train #** | **Test #** | **Dice_network** | **Dice_post** | **Dice_gain_post** | **Dice_P_value** | **Jaccard_network** | **Jaccard_post** | **Jacc_gain_post** | **Jacc_P_value** |
| --- | --- | --- | --- | --- | --- | --- | --- | --- | --- | --- | --- |
| **Liver** | database #0 | 172 | 43 | 96.19 ± 2.19 | 96.28 ± 2.16 | 0.09 ± 0.16 | 3.45E-05 | 92.74 ± 3.90 | 92.90 ± 3.85 | 0.16 ± 0.29 | 3.27E-05 |
|  | database #1 | 20 | 5 | 96.60 ± 0.53 | 96.62 ± 0.55 | 0.02 ± 0.04 | 0.8125 | 93.43 ± 0.98 | 93.46 ± 1.02 | 0.03 ± 0.07 | 0.8125 |
|  | database #3 | 104 | 26 | 96.86 ± 1.39 | 96.92 ± 1.32 | 0.06 ± 0.14 | 0.00221 | 93.94 ± 2.57 | 94.05 ± 2.44 | 0.10 ± 0.25 | 0.002029683 |
|  | database #4 | 292 | 73 | 97.60 ± 1.05 | 97.64 ± 1.03 | 0.03 ± 0.12 | 0.004216709 | 95.34 ± 1.96 | 95.40 ± 1.92 | 0.06 ± 0.22 | 0.004365291 |
|  | database #13 | 232 | 58 | 96.71 ± 1.72 | 96.80 ± 1.62 | 0.09 ± 0.17 | 2.37E-07 | 93.67 ± 3.07 | 93.84 ± 2.91 | 0.16 ± 0.30 | 2.37E-07 |
|  | database #15 | 88 | 22 | 96.83 ± 1.81 | 96.86 ± 1.80 | 0.02 ± 0.03 | 0.000875569 | 93.91 ± 3.29 | 93.96 ± 3.28 | 0.05 ± 0.06 | 0.000875569 |
|  | **All databases** | **908** | **227** | 96.93 ± 1.67 | 96.98 ± 1.62 | 0.06 ± 0.14 | 2.01E-17 | 94.08 ± 3.01 | 94.19 ± 2.93 | 0.11 ± 0.25 | 1.88E-17 |
| **Heart** | database #9 | 40 | 10 | 89.93 ± 2.59 | 90.42 ± 2.51 | 0.50 ± 0.48 | 0.01953125 | 81.79 ± 4.27 | 82.61 ± 4.19 | 0.82 ± 0.81 | 0.01953125 |
|  | database #10 | 108 | 27 | 87.38 ± 6.46 | 87.87 ± 6.20 | 0.49 ± 0.88 | 0.001085403 | 78.10 ± 9.28 | 78.84 ± 9.03 | 0.74 ± 1.30 | 0.001181227 |
|  | database #13 | 220 | 55 | 93.97 ± 5.94 | 94.23 ± 5.45 | 0.26 ± 0.91 | 0.034025219 | 89.10 ± 8.66 | 89.50 ± 8.13 | 0.39 ± 1.39 | 0.035464406 |
|  | **All databases** | **368** | **92** | 91.60 ± 6.52 | 91.95 ± 6.13 | 0.35 ± 0.87 | 1.27E-05 | 85.08 ± 9.82 | 85.62 ± 9.38 | 0.54 ± 1.32 | 1.49E-05 |
| **Pancreas** | database #0 | 156 | 39 | 72.30 ± 14.38 | 71.99 ± 16.59 | -0.31 ± 4.93 | 0.040229498 | 58.35 ± 15.94 | 58.49 ± 17.89 | 0.13 ± 4.98 | 0.03889087 |
|  | database #1 | 36 | 9 | 65.63 ± 19.65 | 64.93 ± 20.98 | -0.70 ± 4.09 | 0.3828125 | 51.19 ± 18.23 | 50.20 ± 20.93 | -0.99 ± 3.98 | 0.91015625 |
|  | database #4 | 276 | 69 | 84.40 ± 13.08 | 83.63 ± 14.71 | -0.77 ± 4.66 | 0.009712671 | 74.87 ± 16.81 | 74.12 ± 18.27 | -0.74 ± 5.15 | 0.00905925 |
|  | database #12 | 60 | 15 | 84.93 ± 11.04 | 82.99 ± 13.37 | -1.94 ± 7.46 | 0.761535645 | 75.15 ± 14.96 | 72.81 ± 17.66 | -2.34 ± 8.92 | 0.678771973 |
|  | database #13 | 212 | 53 | 69.34 ± 16.31 | 69.66 ± 17.22 | 0.33 ± 4.53 | 0.000858946 | 55.20 ± 17.54 | 55.82 ± 18.47 | 0.63 ± 5.00 | 0.000622553 |
|  | database #16 | 244 | 61 | 80.12 ± 15.37 | 76.32 ± 16.19 | -3.81 ± 4.71 | 4.34736E-07 | 69.20 ± 18.93 | 64.07 ± 18.58 | -5.13 ± 5.81 | 2.86538E-07 |
|  | **All databases** | **984** | **246** | 77.52 ± 15.99 | 76.24 ± 17.02 | -1.28 ± 5.07 | 0.56285091 | 65.76 ± 19.15 | 64.25 ± 19.78 | -1.50 ± 5.89 | 0.661929314 |
| **Bones** | database #3 | 84 | 21 | 90.97 ± 3.35 | 91.02 ± 3.31 | 0.05 ± 0.10 | 0.053724708 | 83.61 ± 5.66 | 83.68 ± 5.60 | 0.08 ± 0.17 | 0.058186272 |
|  | database #13 | 216 | 54 | 95.29 ± 4.82 | 94.93 ± 4.80 | -0.36 ± 0.31 | 3.77656E-09 | 91.33 ± 7.43 | 90.68 ± 7.41 | -0.65 ± 0.52 | 1.90594E-09 |
|  | **All databases** | **300** | **75** | 94.08 ± 4.84 | 93.84 ± 4.75 | -0.24 ± 0.32 | 3.74783E-08 | 89.17 ± 7.77 | 88.72 ± 7.60 | -0.45 ± 0.56 | 2.11074E-08 |
| **Kidneys** | database #0 | 160 | 40 | 93.50 ± 4.81 | 93.57 ± 5.47 | 0.08 ± 1.28 | 0.459742241 | 88.11 ± 7.48 | 88.33 ± 8.22 | 0.22 ± 1.96 | 0.427756533 |
|  | database #3 | 92 | 23 | 90.64 ± 6.44 | 90.60 ± 6.53 | -0.03 ± 0.21 | 0.223757155 | 83.44 ± 10.07 | 83.40 ± 10.18 | -0.04 ± 0.32 | 0.273543661 |
|  | database #4 | 296 | 74 | 95.37 ± 3.52 | 95.53 ± 3.56 | 0.16 ± 0.51 | 8.60E-09 | 91.33 ± 5.54 | 91.62 ± 5.64 | 0.29 ± 0.86 | 7.10E-09 |
|  | **All databases** | **548** | **137** | 94.03 ± 4.80 | 94.13 ± 5.05 | 0.10 ± 0.79 | 5.10E-05 | 89.07 ± 7.57 | 89.28 ± 7.89 | 0.21 ± 1.24 | 3.23E-05 |
| **Spleen** | database #0 | 152 | 38 | 92.20 ± 11.63 | 93.41 ± 8.66 | 1.21 ± 5.02 | 0.00479201 | 87.12 ± 15.27 | 88.61 ± 12.40 | 1.49 ± 5.10 | 0.004579774 |
|  | database #1 | 28 | 7 | 96.12 ± 0.85 | 96.17 ± 0.80 | 0.06 ± 0.07 | 0.15625 | 92.54 ± 1.57 | 92.64 ± 1.49 | 0.10 ± 0.13 | 0.15625 |
|  | database #4 | 304 | 76 | 97.05 ± 1.92 | 97.00 ± 3.23 | -0.05 ± 3.34 | 0.000439055 | 94.33 ± 3.43 | 94.33 ± 5.09 | -0.00 ± 5.21 | 0.000439055 |
|  | database #13 | 224 | 56 | 91.58 ± 14.32 | 92.09 ± 14.30 | 0.51 ± 1.53 | 0.000147357 | 86.45 ± 15.56 | 87.31 ± 15.47 | 0.86 ± 2.49 | 0.0002747 |
|  | database #17 | 24 | 6 | 95.78 ± 1.40 | 95.79 ± 1.37 | 0.01 ± 0.04 | 1 | 91.94 ± 2.55 | 91.95 ± 2.50 | 0.01 ± 0.07 | 1 |
|  | **All databases** | **732** | **183** | 94.29 ± 9.88 | 94.68 ± 9.28 | 0.39 ± 3.26 | 2.44E-09 | 90.28 ± 11.81 | 90.85 ± 11.16 | 0.57 ± 4.33 | 4.01E-09 |
| **UB** | database #0 | 148 | 37 | 78.88 ± 18.88 | 79.34 ± 18.89 | 0.46 ± 1.46 | 0.017550104 | 68.43 ± 22.12 | 69.09 ± 22.15 | 0.65 ± 1.72 | 0.012210255 |
|  | database #1 | 4 | 1 | 25.73 ± 0.00 | 23.51 ± 0.00 | -2.22 ± 0.00 | 1 | 14.77 ± 0.00 | 13.32 ± 0.00 | -1.45 ± 0.00 | 1 |
|  | database #3 | 88 | 22 | 78.64 ± 21.76 | 79.21 ± 21.55 | 0.57 ± 1.83 | 0.005723187 | 68.98 ± 24.71 | 69.40 ± 25.24 | 0.42 ± 2.84 | 0.020270982 |
|  | database #13 | 224 | 56 | 89.30 ± 12.00 | 89.59 ± 11.93 | 0.28 ± 0.96 | 0.001135215 | 82.25 ± 15.08 | 82.71 ± 15.07 | 0.46 ± 1.40 | 0.000824594 |
|  | **All databases** | **464** | **116** | 83.41 ± 17.99 | 83.78 ± 17.97 | 0.37 ± 1.34 | 4.12E-06 | 74.75 ± 21.24 | 75.24 ± 21.40 | 0.50 ± 1.84 | 6.36E-06 |
| **Esophagus** | database #0 | 160 | 40 | 77.57 ± 9.69 | 76.36 ± 11.99 | -1.21 ± 4.12 | 0.418290922 | 64.26 ± 11.67 | 63.06 ± 13.80 | -1.20 ± 4.27 | 0.394627787 |
|  | database #1 | 24 | 6 | 73.86 ± 5.78 | 74.24 ± 6.01 | 0.38 ± 0.79 | 0.21875 | 58.84 ± 7.42 | 59.35 ± 7.80 | 0.51 ± 0.97 | 0.21875 |
|  | database #9 | 44 | 11 | 71.91 ± 10.42 | 72.46 ± 9.71 | 0.55 ± 2.73 | 0.206054688 | 57.00 ± 11.69 | 57.60 ± 11.26 | 0.60 ± 2.57 | 0.206054688 |
|  | database #10 | 264 | 66 | 72.47 ± 7.98 | 71.63 ± 10.24 | -0.84 ± 4.46 | 0.002335167 | 57.40 ± 9.28 | 56.69 ± 11.38 | -0.71 ± 4.46 | 0.001766205 |
|  | database #13 | 228 | 57 | 71.14 ± 14.10 | 70.16 ± 15.63 | -0.98 ± 4.01 | 0.845661009 | 56.82 ± 15.14 | 55.95 ± 16.35 | -0.87 ± 3.57 | 0.753645814 |
|  | **All databases** | **720** | **180** | 73.20 ± 10.90 | 72.35 ± 12.55 | -0.84 ± 4.07 | 0.010390799 | 58.76 ± 12.27 | 58.01 ± 13.73 | -0.75 ± 3.98 | 0.009761874 |
| **Femur Heads** | database #13 | 220 | 55 | 95.65 ± 2.78 | 95.72 ± 2.79 | 0.07 ± 0.14 | 3.90E-06 | 91.78 ± 4.77 | 91.92 ± 4.79 | 0.14 ± 0.24 | 4.06E-06 |
|  | **All databases** | **220** | **55** | 95.65 ± 2.78 | 95.72 ± 2.79 | 0.07 ± 0.14 | 3.90E-06 | 91.78 ± 4.77 | 91.92 ± 4.79 | 0.14 ± 0.24 | 4.06E-06 |
| **Lungs** | database #0 | 108 | 27 | 97.30 ± 1.40 | 97.39 ± 1.14 | 0.08 ± 0.35 | 0.501138185 | 94.78 ± 2.60 | 94.93 ± 2.15 | 0.15 ± 0.63 | 0.501138185 |
|  | database #1 | 16 | 4 | 97.63 ± 1.07 | 97.62 ± 1.08 | -0.02 ± 0.03 | 0.625 | 95.39 ± 2.04 | 95.36 ± 2.05 | -0.03 ± 0.06 | 0.625 |
|  | database #3 | 92 | 23 | 97.85 ± 0.80 | 97.82 ± 0.82 | -0.03 ± 0.08 | 0.016270915 | 95.81 ± 1.53 | 95.75 ± 1.55 | -0.06 ± 0.14 | 0.017674556 |
|  | database #4 | 200 | 50 | 97.42 ± 1.07 | 96.79 ± 4.28 | -0.63 ± 4.17 | 0.018263773 | 95.00 ± 2.01 | 94.04 ± 6.44 | -0.96 ± 6.15 | 0.017793864 |
|  | database #9 | 32 | 8 | 95.31 ± 3.04 | 95.25 ± 3.12 | -0.06 ± 0.08 | 0.1484375 | 91.17 ± 5.41 | 91.08 ± 5.53 | -0.10 ± 0.14 | 0.1953125 |
|  | database #10 | 200 | 50 | 97.89 ± 0.57 | 97.88 ± 0.58 | -0.01 ± 0.04 | 0.392930388 | 95.88 ± 1.09 | 95.86 ± 1.11 | -0.01 ± 0.08 | 0.398299647 |
|  | database #12 | 52 | 13 | 97.62 ± 0.69 | 94.86 ± 5.53 | -2.76 ± 5.53 | 0.057373047 | 95.36 ± 1.31 | 90.68 ± 9.34 | -4.68 ± 9.33 | 0.057373047 |
|  | database #13 | 204 | 51 | 98.00 ± 0.91 | 97.98 ± 0.92 | -0.02 ± 0.12 | 0.606175252 | 96.09 ± 1.73 | 96.06 ± 1.75 | -0.03 ± 0.22 | 0.606175252 |
|  | database #15 | 72 | 18 | 98.15 ± 0.61 | 97.93 ± 0.92 | -0.23 ± 0.71 | 0.000233229 | 96.38 ± 1.18 | 95.95 ± 1.75 | -0.43 ± 1.34 | 0.000233229 |
|  | database #16 | 148 | 37 | 97.54 ± 0.95 | 97.26 ± 1.28 | -0.29 ± 0.87 | 3.76507E-06 | 95.22 ± 1.80 | 94.69 ± 2.38 | -0.53 ± 1.60 | 3.76507E-06 |
|  | database #17 | 36 | 9 | 97.32 ± 0.65 | 97.23 ± 0.72 | -0.09 ± 0.14 | 0.01171875 | 94.79 ± 1.23 | 94.63 ± 1.37 | -0.17 ± 0.27 | 0.01171875 |
|  | database #18 train | 12 | 3 | 97.31 ± 1.35 | 88.17 ± 7.42 | -9.14 ± 7.98 | 0.5 | 94.79 ± 2.58 | 79.40 ± 12.38 | -15.39 ± 13.50 | 0.5 |
|  | database #18 validation | 28 | 7 | 97.95 ± 0.83 | 80.95 ± 10.26 | -16.99 ± 9.86 | 0.03125 | 95.99 ± 1.59 | 69.13 ± 15.37 | -26.85 ± 14.63 | 0.03125 |
|  | database #18 test | 20 | 5 | 98.09 ± 0.41 | 77.95 ± 11.32 | -20.14 ± 11.33 | 0.0625 | 96.24 ± 0.78 | 65.01 ± 15.50 | -31.23 ± 15.53 | 0.0625 |
|  | database #19 validation | 4 | 1 | 90.06 ± 0.00 | 90.06 ± 0.00 | 0.00 ± 0.00 | 1 | 81.92 ± 0.00 | 0.00 ± 0.00 | -81.92 ± 0.00 | 1 |
|  | database #19 training | 16 | 4 | 97.66 ± 0.52 | 97.66 ± 0.54 | 0.00 ± 0.02 | 1 | 95.44 ± 1.00 | 95.44 ± 1.02 | 0.00 ± 0.04 | 1 |
|  | **All databases** | **1240** | **310** | 97.63 ± 1.18 | 96.56 ± 4.70 | -1.07 ± 4.61 | 5.03027E-13 | 95.39 ± 2.18 | 93.42 ± 9.06 | -1.97 ± 8.46 | 3.47843E-13 |
| **SC** | database #9 | 60 | 15 | 87.73 ± 4.10 | 87.80 ± 4.13 | 0.07 ± 0.21 | 0.359130859 | 78.35 ± 6.23 | 78.46 ± 6.28 | 0.11 ± 0.32 | 0.33026123 |
|  | database #10 | 332 | 83 | 88.72 ± 3.66 | 88.89 ± 3.69 | 0.18 ± 0.48 | 2.38E-07 | 79.90 ± 5.46 | 80.19 ± 5.52 | 0.29 ± 0.76 | 9.81E-08 |
|  | database #13 | 196 | 49 | 91.92 ± 2.18 | 92.10 ± 2.29 | 0.18 ± 0.27 | 5.73E-05 | 85.13 ± 3.65 | 85.44 ± 3.83 | 0.31 ± 0.46 | 4.07E-05 |
|  | **All databases** | **588** | **147** | 89.69 ± 3.65 | 89.85 ± 3.70 | 0.17 ± 0.39 | 8.17E-11 | 81.49 ± 5.63 | 81.76 ± 5.72 | 0.28 ± 0.64 | 2.60E-11 |
| **Aorta** | database #0 | 156 | 39 | 93.43 ± 2.28 | 93.88 ± 1.73 | 0.45 ± 1.05 | 7.01E-06 | 87.76 ± 3.89 | 88.52 ± 3.03 | 0.76 ± 1.72 | 7.01E-06 |
|  | database #1 | 28 | 7 | 90.53 ± 4.44 | 91.67 ± 3.62 | 1.14 ± 2.48 | 0.03125 | 82.94 ± 7.21 | 84.79 ± 5.90 | 1.85 ± 3.94 | 0.03125 |
|  | **All databases** | **184** | **46** | 92.99 ± 2.85 | 93.55 ± 2.22 | 0.56 ± 1.34 | 6.12E-07 | 87.03 ± 4.77 | 87.95 ± 3.77 | 0.93 ± 2.18 | 5.78E-07 |
| **GB** | database #0 | 148 | 37 | 76.21 ± 17.88 | 75.62 ± 18.68 | -0.59 ± 6.31 | 0.875161507 | 64.32 ± 19.98 | 63.16 ± 22.19 | -1.17 ± 7.88 | 0.791772213 |
|  | database #1 | 12 | 3 | 70.37 ± 9.26 | 80.48 ± 9.34 | 10.11 ± 11.77 | 0.5 | 54.81 ± 10.88 | 68.01 ± 12.88 | 13.20 ± 14.40 | 0.5 |
|  | database #13 | 224 | 56 | 80.87 ± 16.42 | 81.70 ± 15.48 | 0.83 ± 3.52 | 0.03838147 | 70.45 ± 19.25 | 70.16 ± 20.34 | -0.30 ± 9.18 | 0.151102853 |
|  | **All databases** | **384** | **96** | 78.74 ± 16.92 | 79.32 ± 16.78 | 0.57 ± 5.35 | 0.095184994 | 67.60 ± 19.56 | 67.39 ± 21.00 | -0.21 ± 9.09 | 0.297653489 |
| **AG** | database #0 | 164 | 41 | 59.40 ± 18.71 | 55.53 ± 17.91 | -3.87 ± 7.38 | 0.000296619 | 44.34 ± 16.31 | 33.78 ± 18.67 | -10.56 ± 14.32 | 9.63175E-06 |
|  | database #1 | 16 | 4 | 45.09 ± 19.71 | 42.97 ± 23.27 | -2.12 ± 4.23 | 1 | 30.65 ± 16.22 | 6.96 ± 8.10 | -23.69 ± 14.17 | 0.125 |
|  | database #13 | 200 | 50 | 51.51 ± 15.33 | 49.11 ± 16.39 | -2.39 ± 4.38 | 0.001039262 | 36.01 ± 13.12 | 29.00 ± 17.47 | -7.01 ± 11.03 | 4.42962E-06 |
|  | **All databases** | **380** | **95** | 54.64 ± 17.40 | 51.62 ± 17.52 | -3.02 ± 5.86 | 5.21551E-07 | 39.38 ± 15.21 | 30.14 ± 18.41 | -9.24 ± 13.01 | 1.83648E-11 |
| **IVC** | **All databases** | 148 | 37 | 80.25 ± 11.92 | 62.95 ± 11.46 | -17.30 ± 11.12 | 1.17419E-06 | 68.45 ± 14.97 | 41.22 ± 13.56 | -27.23 ± 11.94 | 1.14021E-07 |
|  | database #1 | 36 | 9 | 81.87 ± 8.72 | 61.09 ± 11.91 | -20.79 ± 9.28 | 0.0078125 | 70.10 ± 12.05 | 39.94 ± 12.43 | -30.16 ± 7.97 | 0.00390625 |
|  | **All databases** | **184** | **46** | 80.57 ± 11.30 | 62.59 ± 11.44 | -17.98 ± 10.78 | 5.2553E-08 | 68.77 ± 14.33 | 40.97 ± 13.22 | -27.80 ± 11.26 | 3.52295E-09 |
| **Colon** | database #13 | 224 | 56 | 70.61 ± 11.97 | 67.48 ± 15.25 | -3.13 ± 7.05 | 0.166830364 | 55.76 ± 13.20 | 52.16 ± 16.96 | -3.60 ± 8.15 | 0.133380513 |
|  | **All databases** | **224** | **56** | 70.61 ± 11.97 | 67.48 ± 15.25 | -3.13 ± 7.05 | 0.166830364 | 55.76 ± 13.20 | 52.16 ± 16.96 | -3.60 ± 8.15 | 0.133380513 |
| **Rectum** | database #13 | 228 | 57 | 77.90 ± 13.31 | 77.70 ± 13.33 | -0.20 ± 2.44 | 0.063566803 | 65.41 ± 15.13 | 65.14 ± 15.23 | -0.26 ± 3.31 | 0.048338072 |
|  | **All databases** | **228** | **57** | 77.90 ± 13.31 | 77.70 ± 13.33 | -0.20 ± 2.44 | 0.063566803 | 65.41 ± 15.13 | 65.14 ± 15.23 | -0.26 ± 3.31 | 0.048338072 |
| **SI** | database #1 | 8 | 2 | 17.18 ± 14.83 | 15.96 ± 16.76 | -1.21 ± 1.93 | 1 | 9.76 ± 8.90 | 9.13 ± 9.94 | -0.63 ± 1.04 | 1 |
|  | database #13 | 220 | 55 | 72.74 ± 9.78 | 73.47 ± 9.76 | 0.73 ± 0.91 | 8.19E-09 | 58.04 ± 11.71 | 58.40 ± 13.42 | 0.35 ± 4.41 | 4.26E-08 |
|  | **All databases** | **228** | **57** | 70.79 ± 14.23 | 71.45 ± 14.52 | 0.66 ± 1.00 | 5.07E-08 | 56.35 ± 14.63 | 56.67 ± 16.10 | 0.32 ± 4.34 | 1.47E-07 |
| **Stomach** | database #0 | 172 | 43 | 86.18 ± 10.31 | 75.52 ± 10.96 | -10.66 ± 10.15 | 6.71152E-07 | 76.90 ± 13.43 | 59.26 ± 16.67 | -17.64 ± 15.56 | 1.70933E-07 |
|  | database #1 | 12 | 3 | 72.56 ± 25.54 | 61.17 ± 30.78 | -11.39 ± 9.89 | 0.5 | 60.88 ± 29.57 | 27.42 ± 29.01 | -33.47 ± 26.18 | 0.25 |
|  | database #13 | 224 | 56 | 86.99 ± 9.86 | 75.59 ± 11.61 | -11.40 ± 12.60 | 1.69149E-07 | 78.16 ± 13.91 | 57.21 ± 18.84 | -20.95 ± 19.39 | 9.32407E-09 |
|  | **All databases** | **408** | **102** | 86.22 ± 10.76 | 75.13 ± 12.17 | -11.09 ± 11.46 | 2.32203E-13 | 77.12 ± 14.36 | 57.20 ± 18.80 | -19.92 ± 18.09 | 2.64761E-15 |
| **Thymus** | database #13 | 108 | 27 | 64.11 ± 23.55 | 65.36 ± 23.57 | 1.25 ± 2.63 | 0.000545135 | 50.89 ± 22.67 | 52.26 ± 22.75 | 1.37 ± 3.01 | 0.001708899 |
|  | **All databases** | **108** | **27** | 64.11 ± 23.55 | 65.36 ± 23.57 | 1.25 ± 2.63 | 0.000545135 | 50.89 ± 22.67 | 52.26 ± 22.75 | 1.37 ± 3.01 | 0.001708899 |
| **Gis** | database #13 | 220 | 55 | 88.88 ± 8.87 | 89.26 ± 8.22 | 0.38 ± 1.44 | 0.000142458 | 80.91 ± 11.79 | 81.40 ± 11.14 | 0.50 ± 1.66 | 0.000142458 |
|  | **All databases** | **220** | **55** | 88.88 ± 8.87 | 89.26 ± 8.22 | 0.38 ± 1.44 | 0.000142458 | 80.91 ± 11.79 | 81.40 ± 11.14 | 0.50 ± 1.66 | 0.000142458 |
| **Eyes** | database #2 | 4 | 1 | 83.32 ± 0.00 | 83.34 ± 0.00 | 0.02 ± 0.00 | 1 | 71.41 ± 0.00 | 71.44 ± 0.00 | 0.04 ± 0.00 | 1 |
|  | database #5 | 40 | 10 | 85.33 ± 2.46 | 85.33 ± 2.46 | 0.00 ± 0.00 | 1 | 74.49 ± 3.75 | 0.00 ± 0.00 | -74.49 ± 3.75 | 0.001953125 |
|  | database #8 | 12 | 3 | 90.81 ± 2.76 | 90.82 ± 2.78 | 0.01 ± 0.02 | 1 | 83.24 ± 4.69 | 29.57 ± 51.23 | -53.66 ± 46.54 | 0.5 |
|  | database #11 | 348 | 87 | 91.89 ± 2.32 | 91.89 ± 2.35 | -0.00 ± 0.10 | 0.991373759 | 85.08 ± 3.82 | 28.05 ± 39.87 | -57.03 ± 40.66 | 7.28907E-13 |
|  | **All databases** | **404** | **101** | 91.13 ± 3.13 | 91.13 ± 3.15 | 0.00 ± 0.09 | 0.860004132 | 83.84 ± 5.09 | 25.75 ± 38.89 | -58.09 ± 39.10 | 4.13233E-15 |
| **Brain** | database #2 | 4 | 1 | 95.42 ± 0.00 | 95.42 ± 0.00 | 0.00 ± 0.00 | 1 | 91.24 ± 0.00 | 0.00 ± 0.00 | -91.24 ± 0.00 | 1 |
|  | database #3 | 8 | 2 | 97.80 ± 1.09 | 97.82 ± 1.03 | 0.02 ± 0.05 | 1 | 95.71 ± 2.08 | 95.75 ± 1.98 | 0.04 ± 0.10 | 1 |
|  | database #5 | 40 | 10 | 98.15 ± 0.42 | 98.15 ± 0.42 | 0.00 ± 0.00 | 0.5 | 96.37 ± 0.80 | 19.36 ± 40.81 | -77.01 ± 40.60 | 0.009765625 |
|  | database #7 | 52 | 13 | 98.31 ± 0.44 | 97.91 ± 1.69 | -0.40 ± 1.46 | 0.84375 | 96.68 ± 0.85 | 62.02 ± 44.29 | -34.66 ± 44.63 | 0.039794922 |
|  | database #8 | 32 | 8 | 94.00 ± 2.84 | 89.40 ± 13.77 | -4.59 ± 13.00 | 1 | 88.79 ± 5.09 | 4.86 ± 13.76 | -83.93 ± 15.50 | 0.0078125 |
|  | database #11 | 132 | 33 | 97.66 ± 0.52 | 97.53 ± 0.80 | -0.13 ± 0.78 | 0.556640625 | 95.43 ± 0.99 | 29.13 ± 44.03 | -66.30 ± 43.96 | 1.06067E-05 |
|  | database #14 | 8 | 2 | 97.10 ± 1.29 | 97.08 ± 1.32 | -0.02 ± 0.03 | 1 | 94.38 ± 2.44 | 46.29 ± 65.46 | -48.09 ± 67.91 | 0.5 |
|  | **All databases** | **276** | **69** | 97.39 ± 1.66 | 96.72 ± 5.26 | -0.67 ± 4.48 | 0.909011307 | 94.95 ± 3.03 | 33.10 ± 44.58 | -61.85 ± 44.10 | 2.28764E-10 |

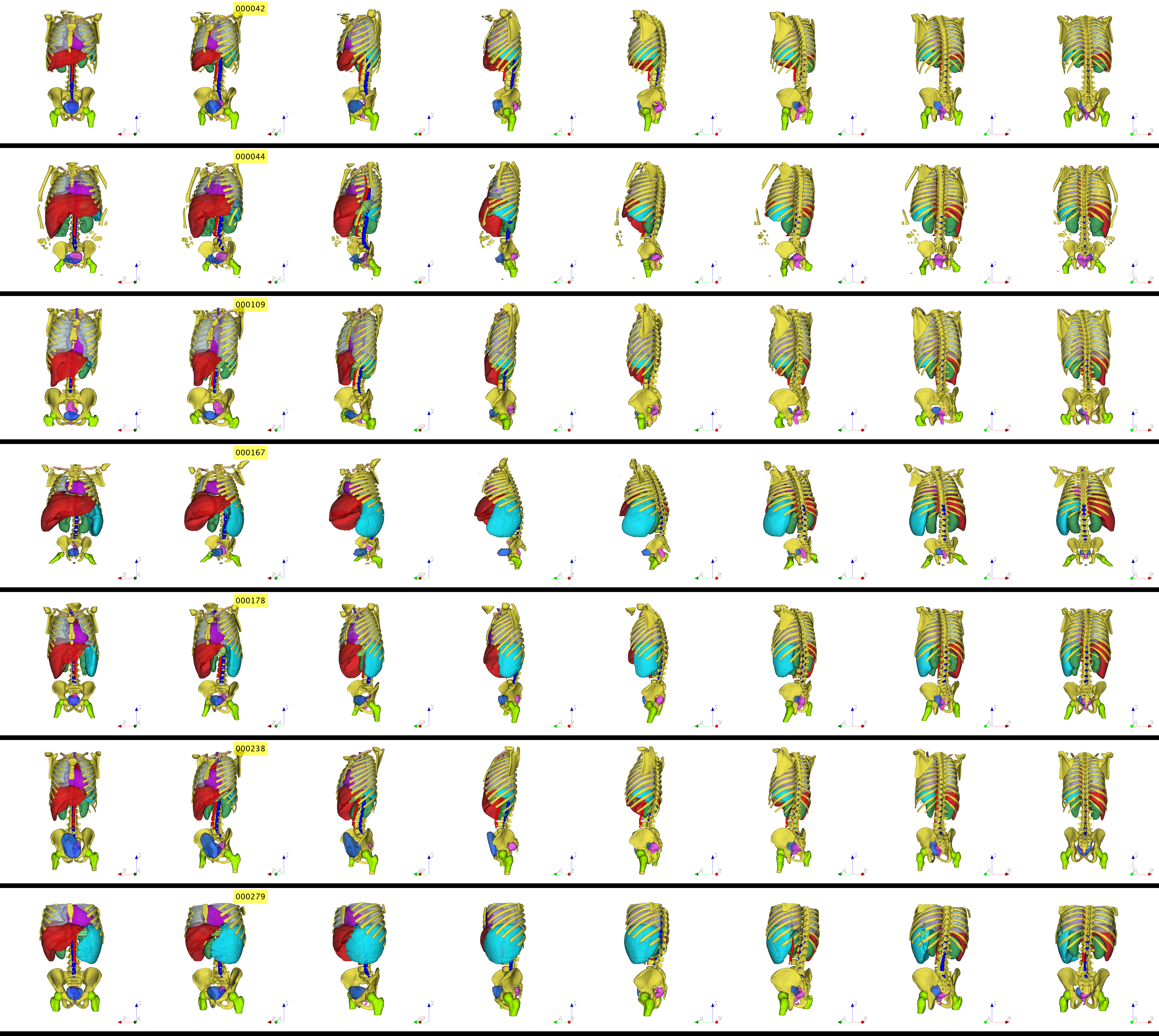

**Supplementary Figure 1.** 3D visualization of segmentation masks of different organs showing: the kidneys (dark green), femoral heads (lime), bones (yellow), liver (dark red), aorta (light red), spleen (cyan), heart (purple), stomach (light green), spinal cord (dark blue), urinary bladder (light blue), and the rectum (pink).
